# Could behaviour change techniques be used to address under-recognition of work-related asthma in primary care? A systematic review

**DOI:** 10.1101/2024.02.22.24303183

**Authors:** GI Walters, H Foley, CC Huntley, A Naveed, K Nettleton, C Reilly, M Thomas, C Walker, K Wheeler

## Abstract

**Introduction:** Work-related asthma (WRA) is prevalent yet under-recognized in UK primary care. The aim of this systematic review was to identify behaviour change interventions (BCI) intended for use in a primary care setting to identify any chronic disease, that may be used in the context of WRA. The study was registered on the PROPSPERO database (19/04/2023; CRD42023418316) and received no funding.

**Methods:** We searched CCRCT, Embase, PsychINFO and Ovid MEDLINE databases (1^st^ January 1946 - 6^th^ March 2023) for any observational or experimental study which described the development or evaluation (or both) of a BCI for case finding any chronic disease in a primary care setting, aimed at either healthcare professionals or patients or both. We included case reports, series and conference abstracts, and excluded existing reviews and protocols, and abstracts not in English. Abstracts and subsequent full text articles were assessed by two blinded, independent reviewers, and disagreement resolved by consensus. The primary author undertook quality assessments for a variety of methodologies, with quality control by a second reviewer. We undertook narrative synthesis for a variety of outcomes of usability and effectiveness, and for BCI development.

**Results:** 18 studies (14 papers and 4 conference abstracts) were included following full-text review, from an initial literature search yielding n=768 citations for screening, of which there were 3 randomised control trials, 1 uncontrolled experimental study, 4 primarily qualitative studies and 10 studies employing recognized multi-step BC methodologies. Quality varied depending upon the methodology used. None of the studies were concerned with identification of asthma. BCIs had been developed for facilitating screening programmes (5), implementing guidelines (5) and individual case finding (8). Six studies measured effectiveness, in terms of screening adherence rates, pre- and post-intervention competency, satisfaction and usability, for clinicians, though none measured diagnostic rates.

**Discussion:** Single and multi-component BCIs have been developed to aid identification of chronic diseases, though not asthma or work-related asthma specifically. Development for the majority has used BC methodologies that involve gathering data from a range of sources, and develop content specific to defined at-risk populations. Nevertheless, such methodologies could be used similarly to develop a BCI for WRA in primary care settings.

## Introduction

Work-related asthma (WRA) includes asthma causes by exposures at work (occupational asthma) and existing asthma worsening by conditions at work (work-exacerbated asthma). The impetus for this study derives from our clinical experience of WRA as a markedly under-recognized chronic condition. If diagnosis is delayed or missed, affected individuals may suffer poor outcomes in terms of respiratory health and employment [Barber et al., 2022; HSE, 2022], and the societal impacts of healthcare-related costs and productivity losses are significant, with desktop estimates of social cost for the UK in the region of £1 billion per decade [Ayres et al., 2011]. Existing guidelines recommend that attending healthcare professionals (HCP) ask adult patients with new, reactivated or worsening asthma symptoms about the nature of their job, about any relationship of their asthma symptoms with work, and seek expert advice when a relationship is demonstrated [Barber et al., 2022; NICE, 2023]. However, in the UK and elsewhere, it is evident that these guidelines and their previous iterations have not been implemented successfully in a variety of settings, including primary care [Walters et al., 2012; de Bono and Hudsmith, 1999; Fishwick et al., 2007]. In primary care, barriers to diagnosis for patients include poor understanding of asthma and work context, fear of job and economic loss, and the assumption of insolubility [Walters et al., 2015; Bradshaw et al., 2007]. For primary care HCPs barriers include inadequate training and clinical experience, beliefs and perceptions about disease occurrence, pressures on specialist referral, lack of continuity of care, and time constraints [Walters et al., 2021].

Behaviour change interventions (BCI), including education and training aimed at modifying clinical behaviours, audit and feedback, or enablement of collaborative team-based approaches, are broadly effective in changing the individual or collective practice of primary care HCPs [Chauhan et al., 2017]. Additionally, similar approaches may be engaged to influence the health-seeking behaviour of patients. A simple or complex BCI intended to increase early identification of WRA in primary care could be developed and evaluated for clinical effectiveness. An important step, and the rationale for this review, would be to see whether such interventions already exist, or whether similar initiatives from other chronic disease areas could be adapted for this purpose.

### Review questions

(1) Have BCIs aimed at modifying health-seeking or clinical behaviour been developed and/or evaluated for identifying (1) WRA or (2) any chronic disease, in primary care settings?

(2) Are such BCIs effective in aiding identification of such chronic diseases?

## Methods

This systematic review has been reported according to The Preferred Reporting Items for Systematic reviews and Meta-Analyses (PRISMA) statement, 2020 update and PRISMA 2020 for abstracts checklist [Page et al., 2021]. The protocol was registered on the international PROPSERO database of systematic reviews on 19^th^ April 2023 (Registration number: CRD42023418316).

### Literature search

The literature search was undertaken via institutional access to the Ovid interface (Ovid technologies, New York City, United States) and included the following databases: Cochrane Central Register of Controlled Trials (to 6^th^ March 2023), Embase (<1974-March 6^th^ 2023), Ovid MEDLINE (1946-March 6^th^ 2023) and APA PsycINFO (<1967-March 6^th^ 2023). Search terms combined synonyms for three elements: BCIs, diagnosis and primary care; the search strategy is shown in Supplementary Table 1. Reference lists from studies meeting the inclusion criteria were searched to identify any missed relevant studies.

### Inclusion criteria

Any study examining either the development, effectiveness, or both, of the BCI(s) in question, to include qualitative, observational, or experimental studies in biomedical and social science journals. These may have included case reports, case series, published conference abstracts.

### Exclusion Criteria

(1) Books, book chapters, theses, dissertations, systematic or narrative reviews, opinion pieces, protocols (unless a protocol for an effectiveness trial also describes the development of the intervention)

(2) Abstracts not in English

(3) Non-human research (eg. laboratory studies)

### Condition being studied

Any chronic disease, defined here as a disease of any bodily system likely to remain active or require treatment for more than 1 year, and that affects day-to-day functioning, requires medical treatment, or negatively impacts life expectancy. Common examples might include hypertension, diabetes mellitus, epilepsy, depression, multiple sclerosis, coeliac disease (not exhaustive). This definition also includes solid-body and haematological cancers.

### Populations being studied

(1) Studies undertaken in any primary care population, with no geographical restriction

AND

(2) Studies in which the target groups are either patients or primary care HCPs, or both

### Interventions being studied

Any tool, intervention or initiative described as a ‘behaviour change intervention’ or in such terms, designed to change the health seeking behaviour of patients or the clinical behaviour of primary care HCPs, to aid diagnosis of any chronic disease. This did not include BCIs aimed at uptake of lifestyle modifications or treatments to prevent onset of disease (primary prevention and health promotion). BCIs aimed at increasing participation in screening programmes were considered for inclusion, though studies which described development or evaluation of screening programmes *per se*, were excluded from this review.

### Comparator groups being studied

For some relevant and included study designs, no comparator group will have been evident (for example, studies describing the development of BCIs may have been mixed-methods, qualitative, protocols for subsequent trials, or case series). Studies evaluating the effectiveness of BCIs may have included randomised controlled trials, where the comparator groups were either existing standards of care or head-to-head comparisons between novel interventions; or may have been uncontrolled trials.

### Primary outcomes

(1) Related to development of BCI: theoretical framework used (if any), method(s) of development, construct, and function of BCI, any validation undertaken.

(2) Related to clinical effectiveness of BCI: any effect measure (for example, risk ratios, prevalence ratios, incidence rate ratios) depending upon the experimental study design.

### Data selection

Results of literature search were exported in to the Rayyan web-based application (Qatar Computing Research Institute, Doha, Qatar) for abstract screening, and automatically de-duplicated. Each abstract was screened independently and blinded by two reviewers, and any abstract included by at least one reviewer was shortlisted for full-text review. Full-text review was carried out independently and blinded by two reviewers, and a 3^rd^ independent review and discussion with consensus between reviewers undertaken where disagreement occurred. Primary reasons for exclusion were documented according to the categories shown in Supplementary Table 2.

### Data gathering

The following data were gathered by the primary author according to a standardised template (detailed in Supplementary Table 3): author(s), year, region, rationale, study design, population, disease entity, BCI, theoretical framework used, construct and function of BCI, any validation undertaken; for RCTs, comparator groups, effect measures and statistical significance were also collected. The data from a small sample (>10%, 3 studies) were checked for accuracy by a 2^nd^ reviewer.

### Risk of bias/quality assessment

For each included full-text article, the primary author and a second reviewer undertook blinded quality assessments (Supplementary Tables 4-7). Where the researchers had employed a conventional study design as the primary methodology (ie. qualitative, RCT, uncontrolled trial), a standardised Joanna Briggs Institute critical appraisal tool was used to assess quality and risk of bias [Barker et al., 2023; Lockwood et al., 2015; Tufanaru et al., 2017]. These were chosen in order to assess quality and risk of bias at study and outcome levels, for a variety of study designs. Where methodology non-traditional (ie. a multi-step or multi-component BCI methodology) then a narrative appraisal was undertaken, and any factors that would reduce confidence in the study design documented. Any disagreements were moderated by consensus.

### Data synthesis

Included studies were grouped primarily by the intended scope of the developed BCI (ie. facilitation of screening, uptake of existing guidelines, individual case finding) and summary of overall study design provided, in terms of geographical reach, aim (BCI development, evaluation or both), target group (patients, HCPs, both) and disease area. No quantitative synthesis or meta-analysis was planned, since pilot searches had revealed heterogeneous study designs, target populations and high likelihood of multi-component studies being included. Studies were considered to have BC methodology if multi-step (>1) components were included to develop or refine BCIs; any underlying theoretical framework was assessed.

Data was tabulated where possible, grouped by variables ie. study design (eg. co-creation, qualitative), disease, target group (patients and/or healthcare professionals), narrative description of function, construct, and for effectiveness studies: summaries of intervention and comparator groups, main outcomes and effect sizes with significance and confidence intervals. An accompanying narrative synthesis of quantitative data was undertaken. It was also intended to make recommendations for use of existing BCIs in the context of WRA.

## Results

From the initial database literature searches, n=768 discrete citations were included for abstract screening following automated software de-duplication (see PRISMA flow chart in Figure 1). Seventy-one articles (9%) were shortlisted by at least one abstract reviewer and included for full-text review. On full-text review, agreement was found between both reviewers in 55/71 articles (77%; including n=16 for inclusion n=39 for exclusion), with no further studies added after 3^rd^ review and moderation of disagreement; 2 additional articles identified through screening of reference lists by the lead author were included. Thus n=18 articles were included in the subsequent evidence synthesis, comprising 14 full papers and 4 conference abstracts. Reasons for exclusion at full-text review stage were as follows: wrong outcome=25, wrong population=14, wrong study design=15. A list of the excluded studies at full-text review is available from the authors on request. Where conference abstracts have been included, a search for a subsequent full paper has been undertaken, and none of these had been peer-reviewed and published at the time of writing.

**Figure 1.**
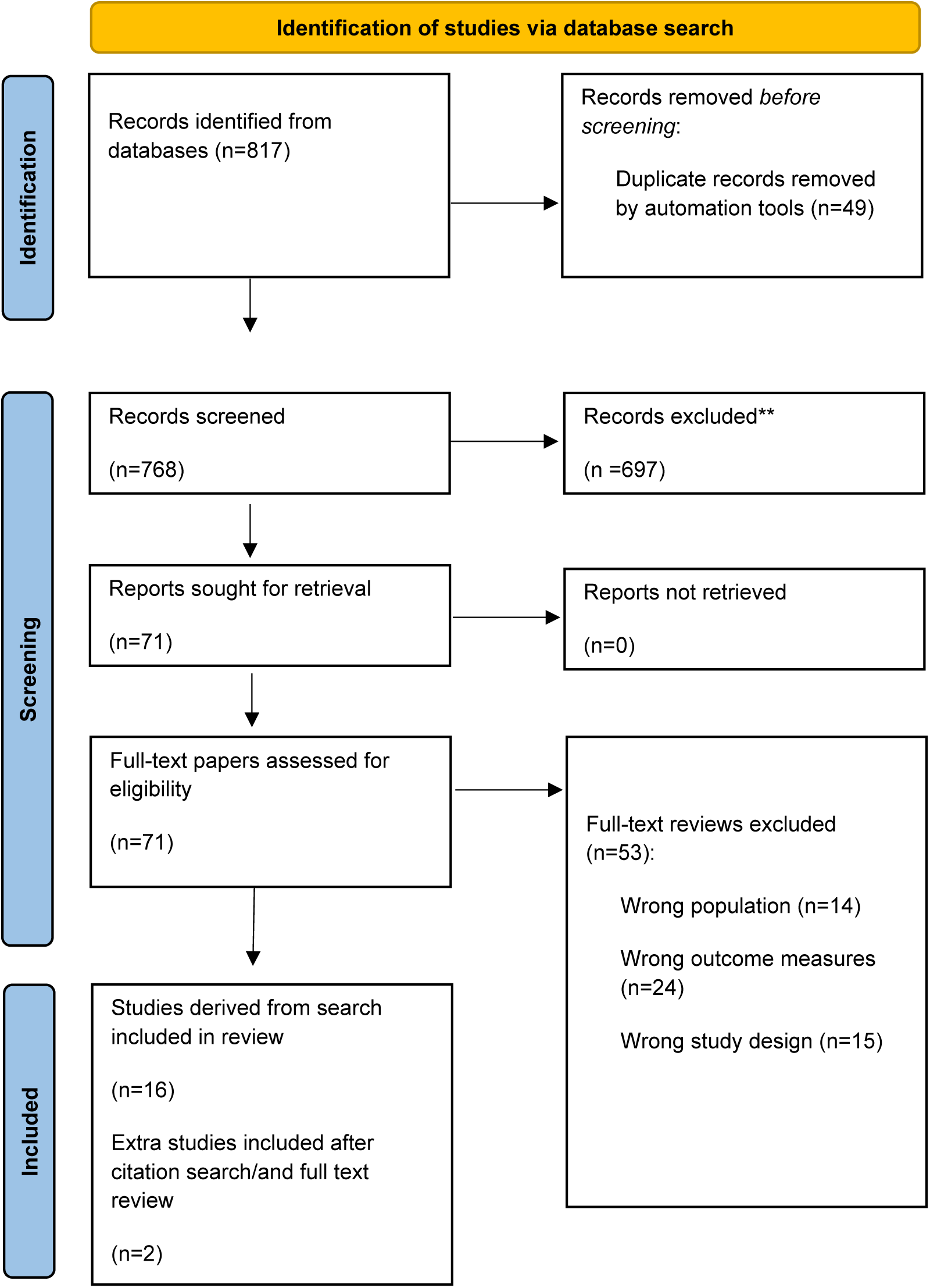
PRISMA study flow chart after Page et al. [2021].

### Quality and risk of bias assessments

Quality and risk of bias assessments are shown in Supplementary Tables 4-7. There were 3 RCTs, 1 quasi-RCT, 4 studies employing predominantly qualitative methodology, and 10 studies describing BCI development using behaviour change (BC) methodology. Included RCTs lacked methodological detail on selection and allocation of participants or sites, in particular the process of randomisation, and on blinding of those evaluating outcomes to allocation; one of these articles [Kronish et al., 2022] was a conference abstract, so many methodological details were absent. The two full-paper RCTs were published in subject-specific journals, so may have lacked the rigorous methodological detail required by a clinical trials journal. The trial with pre- and post-intervention outcome measurements, authored by Porcheret et al. [2018], was nested in a larger RCT described elsewhere, but recruitment and dropout are described in detail. One qualitative study was a conference abstract; methods were described sufficiently to use the correct assessment tool, but lacked detail relating to reflexivity, participant demographics, and ethical permissions. The other 3 studies had full and detailed methods sections.

Critical appraisal of 10 studies employing multi-step BC methodologies revealed inter-study variation, with no single standardised methodology or framework employed. Most commonly the theoretical domains framework (TDF) was used to establish barriers and enablers, with many of these studies applying the behaviour change wheel (BCW) to map barriers and enablers to intervention functions, behaviour change techniques (BCT) and intervention components, and APEASE criteria for final selection [Smits et al., 2018; Jinks et al., 2015; Bravington et al., 2022; Moise et al., 2020; Porcheret et al., 2014; Riordan et al., 2020; Toh et al., 2016, Tuot et al., 2020]. Two studies employed the Medical Research Council (MRC) complex interventions framework [Campbell et al., 2000; Campbell et al., 2007; Lester et al., 2005; Smith et al., 2012]. There were methodological differences between studies for establishing barriers and enablers, identifying and selecting BCTs, and no standardised method for reporting multi-step methods; indeed, due to the need to report multiple work packages in BC methodology studies, the qualitative elements for example, lacked the reporting rigour of papers where qualitative analysis was the sole method used. In some BC methodology papers, individual work packages had been described in detail in separate citations, signposted from the paper [Larkey et al., 2015; Smits et al., 2018; Moise et al., 2020].

### Narrative synthesis

Study characteristics for all included articles are shown in Table 1, grouped by scope of BCI, and displayed with primary methodology, study population, intervention group, comparators, and relevant outcomes.

**Table 1.** Summary of all included studies; ^1^conference abstract. BC=behaviour change; BCI=behaviour change intervention; BCT=behaviour change technique; BCW=behaviour change wheel; GP=general practitioner; HCP=healthcare professional; Medical Research Council=MRC; RCT=randomised controlled trial; SR=systematic review; TDF=theoretical domains framework.

| Scope of BCI | Authors (year) | Region | Stage of implementation | Population studied |  | Primary methodology | Intervention | Control/comparator | Outcome measures |
| --- | --- | --- | --- | --- | --- | --- | --- | --- | --- |
|  |  |  |  | Target group | Disease area |  |  |  |  |
| Facilitation and participation in screening programmes | Bravington et al. (2022) | UK | Development | Patients: women aged >50 years, GPs and practice nurses | Cervical cancer | Multi-step BC methodology, using re-analysis of existing qualitative data according to TDF, design of intervention content and mode of delivery via focus groups | Service-user leaflet (patients) and a video animation for practitioner training (HCPs) | n/a | Barriers and enablers of screening adherence, final intervention |
|  | Kronish et al. (2022)<sup>1</sup> | United States | Development; effectiveness | Doctors, practice nurses, patients | Hypertension | Multi-step BCW methodology and subsequent cluster (practice level) RCT | Multi-component BCI comprising education and training for doctors and nurses on screening, access and ordering of ambulatory/home blood pressure monitoring, and patient information | Standard care | Proportion of eligible patients undertaking ambulatory or home blood pressure monitoring |
|  | Larkey et al. (2015) | United States | Development (described in detail elsewhere); effectiveness | Patients of low income aged 50-70 and minority ethnic groups | Colorectal cancer | Head-to-head RCT of two BCIs | Storytelling: a video created from personal stories composited into a drama about colorectal screening | Personal cancer risk instrument based upon Harvard Cancer Risk Index | Proportion of patients screened |
|  | Riordan et al. (2020) | Republic of Ireland | Development | Patients with type 2 diabetes mellitus, and primary care HCPs | Diabetic retinopathy | Multi-step BCW methodology | Multi-component BCI from a range of identified BCTs. Application of APEASE criteria for selection | n/a | Barriers and enablers, final intervention selection |
|  | Rubinstein et al. (2011) | United States | Effectiveness | Patients aged 35-65 | Cancers, unspecified | Cluster (practice-level) RCT | Interactive, internet-based risk assessment tool that collects family history for common cancers, stratifies personal risk, and provides tailored prevention messages | Standard care | Change in screening adherence rates, at distinct levels of personal risk |
| Implementation of existing guidelines | Kredo et al. (2018) | South Africa | Development | Practice nurses and allied health professionals | Chronic diseases, unspecified | Primarily qualitative, using BCW theory and thematic analysis | n/a | n/a | Barriers and enablers of guideline use; interventions for potential future use identified and characterised |
|  | Leather et al. (2022) | UK | Development | GPs | Self-harm (poisoning or injury) | Primarily qualitative, using BCW theory and thematic analysis | n/a | n/a | Barriers and enablers of guideline use; intervention functions and BCTs for potential future use identified and characterised |
|  | Moise et al. (2020) | United States | Development | GPs and patients | Hypertension | Multi-step BCW methodology | Multi-component BCI from a range of identified BCTs | n/a | Barriers and enablers of guidelines use; feasibility and acceptability |
|  | Porcheret et al. (2014) | UK | Development | GPs enrolled in trial described in (Porcheret et al., 2018) | Osteoarthritis | Multi-step BC methodology underpinned by 4 theoretical models (implementation of change, TDF, adult learning and BCI selection model (after Michie et al., 2008) | Workshops for GPs incorporating multi-component BCI | n/a | Content and function according to TDF; workshop schedules; identification of methods and measures to evaluate workshops |
|  | Porcheret et al. (2018) | UK | Effectiveness | GPs | Osteoarthritis | Uncontrolled experimental study nested within a larger RCT. Pre- and post-intervention video-simulated consultations | Workshops for GPs incorporating multi-component BCI | n/a | Change in competency (number of tasks undertaken per GP) and task delivery (number of GPs undertaking a specific task) scores |
| Individual case-finding, other | Jinks et al. (2015)<sup>1</sup> | UK | Development | Practice nurses | Chronic diseases, unspecified | Multi-step BCW methodology and co-design, including evidence synthesis (nature unclear), community of practice and focus groups | A multi-component BCI: enhanced review consultation including tools for case-finding | n/a | Barriers and enablers of case identification, training needs identified |
|  | Lester et al. (2005) | UK | Development; effectiveness | GPs | Episodic psychosis | MRC complex interventions framework, including theoretical phase (SR) and modelling phase (focus groups and training needs analysis); uncontrolled post-intervention survey | Educational-based BCI comprising video-based consultation role-plays, GP- and service user-led discussion | n/a | Identified training needs; self-reported attitude, awareness, knowledge, and satisfaction amongst participating GPs following intervention initial and booster sessions |
|  | Payne and Hysong (2014)<sup>1</sup> | United States | Development | GPs | Chronic diseases, unspecified | Qualitative interview and deductive analysis | Framework for improving acceptance of physician performance feedback, intended to reduce diagnostic error | n/a | Barriers and enablers of accepting performance feedback amongst GPs |
|  | Smith et al. (2012) | UK | Development | Patients | Lung cancer | MRC complex interventions framework, including SR, HCP-group BCI design, thematic analysis of qualitative interviews with patients and HCP focus groups | A detailed self-help manual and extended consultation with a trained nurse, and development of specific action plans (prompts), aimed at reducing time to presentation with GP | n/a | Barriers and enablers of case identification; final intervention selection |
|  | Smits et al. (2018) | UK | Refinement of existing intervention | Patients aged ≥40 living in socio-economically deprived communities | Cancers, unspecified | Multi-step BCW methodology | A touchscreen questionnaire delivered face-to-face by a trained lay advisor, intended to raise awareness of cancer risk and encourage health-seeking behaviour. Application of APEASE criteria for selection | n/a | Barriers and enablers of use. Final intervention selection |
|  | Toh et al. (2016)<sup>1</sup> | Malaysia | Development | Community pharmacists; GPs | Osteoporosis | Multi-step BCW methodology | Multi-component BCI: pharmacist assessment of personal risk, educational session, restructuring and systematic introduction within clinical practice | n/a | Barriers to case identification; final intervention selection |
|  | Tuot et al., (2020) | United States | Development | GPs and patients | Chronic kidney disease | Multi-step BCW methodology | Patient-facing online risk calculator and a clinician-facing clinical practice toolkit | n/a | Barriers and facilitators to diagnosis; final intervention selection |
|  | Tuot et al., (2022) | United States | Effectiveness | GPs and patients | Chronic kidney disease | Qualitative: thematic analysis of think-aloud protocols with GPs and patients | Patient-facing online risk calculator and a clinician-facing clinical practice toolkit (from Tuot et al., 2020) | n/a | Barriers and enablers of use (usability) |

#### (1) Facilitation of screening programmes

Three studies evaluated effectiveness of BCIs in facilitating a screening programme. Rubenstein et al. [2011] undertook a practice cluster RCT of an existing patient-facing internet-based risk assessment tool for stratifying personal risk and providing tailored messages. Absolute screening adherence increased in both intervention and control groups (standard care) at all levels of personal risk, though there were no significant differences between the two groups (unadjusted or adjusted for risk); the authors noted high baseline screening rates in the sample population. Kronish et al. [2022] developed a multi-component BCI (clinician, nursing and patient education and information) for hypertension screening using BC methodology, which was then evaluated by practice cluster RCT against standard care. There was a significant relative increase in ordering and completion of ambulatory and home blood pressure monitoring in the intervention group (0.5% to 4%; p<0.001 vs. 3.1% to 2.8%; p=0.66 for ordering and 0.5% to 3.0%; p<.001 vs. 2.2% to 2.0%, p=0.76 for completion) though absolute adherence levels remained low. Larkey et al. [2015] undertook a RCT of BCIs to encourage colorectal cancer screening adherence in low income and minority ethnic groups. Head-to-head comparison of ‘storytelling’ (a video created from personal stories composited into a drama) versus personal risk control (a risk assessment tool based upon the Harvard Cancer Risk Index) found no significant differences in screening adherence (37% and 42% screened respectively).

Two studies described development of BCIs using methodologies underpinned by the TDF, aimed at increasing screening adherence; these were for cervical cancer [Bravington et al., 2022] comprising a patient leaflet and video animation for HCP training, and diabetic retinopathy [Riordan et al., 2020] incorporating a range of BC techniques (audit/feedback, electronic prompts targeting HCPs, HCP-endorsed reminders (face-to-face, by phone and letter), and patient leaflet). Riordan et al. [2020] employed a multi-step BCW methodology and measured acceptability and feasibility of their BC techniques via stakeholder consensus groups, refining the final BCI by applying APEASE criteria [Michie et al., 2014].

#### (2) Implementation of diagnostic guidelines

Two authors undertook qualitative interview studies, analysed using the TDF framework, to identify barriers and enablers to adoption of clinical practice guidelines by HCPs, for any chronic disease [Kredo et al., 2018] and specifically for self-harm [Leather et al., 2022]. Both studies mapped findings to BCI functions using COM-B/BCW matrices, and identified multiple BCI groups of relevance for clinical practice guideline adherence (including feedback and monitoring, shaping knowledge, natural consequences, associations, repetition and substitutions, antecedents, goals and planning, and self-belief; Leather et al., 2022), stopping short of selection. Kredo et al. specified a number of BCIs suitable for further evaluation including design improvements, accessibility, digital formatting, and patient engagement materials.

Other authors employed multi-step BC methodology; Moise et al. [2020] developed and refined a multi-component BCI aimed at increasing uptake of hypertension guidelines by patients and HCPs, comprising exploration of barriers via qualitative focus groups, mapping of barriers to functions using the BCW, and feasibility evaluation through stakeholder interviews. Porcheret et al. [2014] drew on four theoretical frameworks: implementation of change model, TDF, theoretical mapping of behavioural determinants to behaviour change techniques (BCT; after Michie et al., 2008) and principles of adult learning, in order to develop an enhanced consultation for identifying osteoarthritis through implementation of UK national guidelines. They developed and refined workshops for General Practice that incorporated BCIs developed via (i) HCP consensus discussion, (ii) GP focus groups and (iii) review of known enablers and barriers to innovation implementation; 10 BC techniques were incorporated including information provision, skills rehearsal, and persuasive communication. A subsequent effectiveness study by the same group [Porcheret et al., 2018] described an uncontrolled trial of the final workshop, with GP competency measured through review of a video-recorded simulated consultation, before and after the intervention; competency score increased significantly at two time points, one month (p=0.001) and five months (p=0.001) after the workshop.

#### (3) Individual case-finding

The majority of papers within this scope (n=6) used multi-step BC methodologies and describe development of BCIs to aid individual case finding in: unspecified long terms conditions [Jinks et al., 2015], first episode psychosis [Lester et al., 2005], lung cancer [Smith et al., 2012], unspecified cancers [Smits et al., 2018] and chronic kidney disease [Tuot et al., 2020]. Four studies used multi-step BCW frameworks; Jinks et al. [2015] developed their multi-component enhanced consultation, which included tools for HCPs for case finding (not described in any further detail in conference abstract) using the TDF/COM-B framework, with steps comprising evidence synthesis, community of practice via stakeholder workshops, and thematic analysis of focus groups to assess training needs. Smits et al., [2018] also employed the TDF/BCW framework to refine content and delivery of an existing BCI, ‘The health check’, a touchscreen questionnaire delivered face-to-face by a trained lay advisor, aiming to raise awareness of cancer risk and encourage health-seeking behaviour in the presence of symptoms, and targeting specifically patients ≥40 years old living in socioeconomically deprived communities. The authors also applied APEASE criteria to make judgements on the most appropriate BCI functions for their objectives. Tuot et al. [2020] developed a patient-facing online risk calculator (brief questionnaire, risk assessment, self-management tools) and a clinician-facing ‘Clinical Practice Toolkit’ platform for identifying chronic kidney disease in at-risk populations. The clinician toolkit included a population-risk identifier, education about detection and management, patient health literacy assessment, and quality improvement intervention suggestions. The authors used a TDF/BCW framework in a multi-step process which comprised establishing barriers and enablers to diagnosis (eg. awareness of guidelines, availability of data analytics) from qualitative data, and mapping these to intervention functions (in this case: education, persuasion, enablement, and modelling) to develop the BCI. In a subsequent effectiveness (usability) study Tuot et al. [2022] undertook thematic analysis of think-aloud exercises completed with patients and primary care HCPs, for the patient-facing online risk calculator element only. They found that patients and clinicians could easily navigate the tool, though there was tension between promotion of medical versus lay terminology; the authors recommend that their findings inform development of future education materials. Toh et al. [2016] have used thematic analysis of primary care stakeholder interviews (patients, pharmacists, nurses, doctors), subsequently applying the BCW to identify a multi-component BCI for osteoporosis case finding consisting of (i) a pharmacist-led risk assessment for osteoporosis, (ii) an education session, and (iii) restructuring of the current practice which incorporates this intervention into daily clinic practice.

Two studies employed the MRC framework for complex interventions; Lester et al. [2005] developed an educational BCI for GPs consulting on first-episode psychosis, via a multi-step process including a literature review, analysis of data from focus groups, and feedback gained from surveys of GPs users following initial and booster training. The BCI was well received amongst GPs with increase in self-reported knowledge, skills, and attitudes in psychotic illness. Smith et al. [2012] used the MRC framework to develop a complex intervention for symptomatic patients at risk of lung cancer, which comprised a detailed self-help manual and extended consultation with a trained research nurse, at which specific action plans were devised. Elements comprised evidence review, multi-disciplinary group design, and thematic analysis of user focus group data.

One study used primarily qualitative methodologies to identify barriers and enablers to BCI for unspecified chronic diseases [Payne and Hysong, 2014]. Payne and Hysong investigated the determinants of acceptability in clinician performance feedback, using deductive analysis from qualitative interviews with GPs to identify barriers (eg. untimely feedback, unrealistic advice, performance based on small sample size) and enablers (eg. sense of competition, showing patient-level data).

Table 2 summarises the 6 studies where full papers were available and that described multi-step BC methodologies using TDF (+/− BCW) frameworks.

**Table 2.**
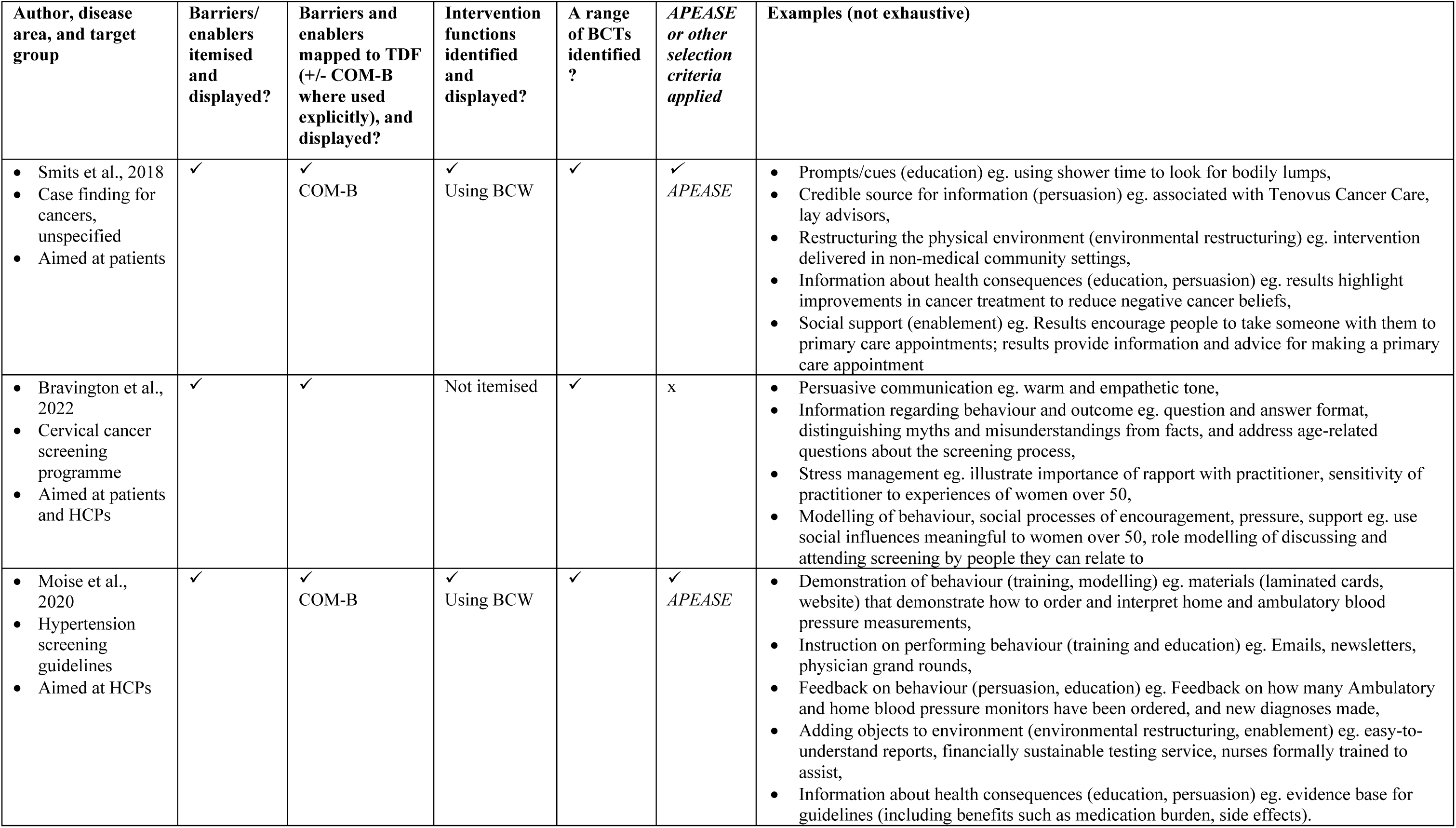
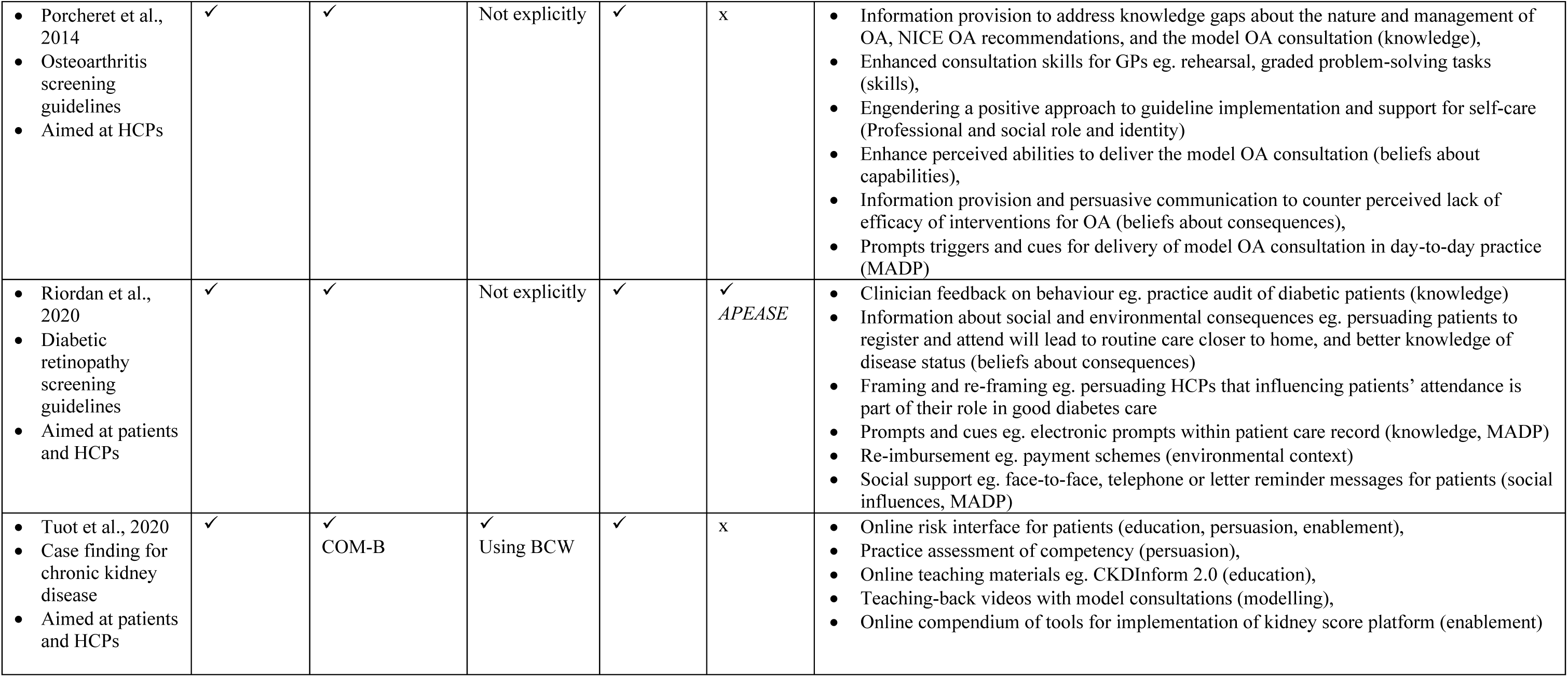
Multi-component BCIs developed using the theoretical domains framework (+/− behaviour change wheel), implied functions, and associated BCTs (full papers only, abstracts excluded). NICE=National Institute for Health and Social Care Excellence; OA=osteoarthritis; HCP=healthcare professional; BCW=behaviour change wheel; TDF=theoretical domains framework; MADP= memory, attention and decision processes.

## Discussion

### Statement of principal findings

This review identified no study describing development of single or multi-component BCIs for the identification of asthma or work-related asthma specifically. Three studies described stages in BCI development for unspecified chronic diseases, that could be employed for asthma, including outlining potential interventions for guideline adherence [Kredo et al., 2018], identifying enablers and barriers to accepting performance feedback for GPs [Payne and Hysong et al. 2014] (conference abstract only), and a multi-component BCI for practice nurses in the format of an enhanced review consultation [Jinks et al., 2015] (conference abstract only). More generally BCI development has focused on adherence to screening programmes, implementation of guidelines and improving individual case finding (eg. through performance feedback, risk assessment and medical education). The majority of studies have drawn on qualitative research with patients and/or HCPs, to define behaviours which require change, usually resulting in identification of multiple BCTs and therefore multi-component BCIs; additionally, a number of theoretical frameworks have been employed, resulting in multi-step development processes, most commonly the TDF (+/− BCW), but also implementation of change theory and the MRC complex interventions framework. Evaluation of BCIs for effectiveness in implementation of guidelines and individual case finding had been limited to outcomes related to diagnostic confidence, competency, improvement in learning, and other self - reported measures amongst HCPs; three RCTs of BCIs aimed to increase screening programme uptake have given both positive and negative results, depending upon background levels of absolute risk.

### Strengths and weaknesses

We have aligned our methods and reporting with the standardised PRISMA checklist and used established quality and risk of bias assessment tools as far as possible. However, no tool or benchmark for assessment of multi-step BC methodologies was available at the time of writing. Often due to the number of work packages involved in a BC methodology study, detail was missing on methods and conduct of individual components, seen most frequently with qualitative methods, though occasionally individual work packages had been published separately. Literature searches were undertaken using search terms which accounted for various spellings of behaviour and acronyms for behaviour change intervention. However, it is possible that such interventions have been missed due to being labelled otherwise.

### Meaning of findings in view of other work

The most encountered framework for research was the TDF, where qualitative methods have been employed to gather data to establish barriers and enablers of behaviour change. This has been frequently undertaken in combination with the more recently developed and simpler COM-B model of behaviour (capability, opportunity, and motivation are the key ingredients of behaviour change), and the BCW approach to intervention design, with demonstrable steps to identification of intervention functions and a range of suitable BCTs. Following a systematic literature review of intervention frameworks, Michie et al. [2011] developed the BCW framework given that no existing framework (including the MRC complex interventions framework) met their criteria of comprehensive coverage of intervention or policy function, coherence, and link to an overarching model of behaviour, including the MRC complex intervention framework. Indeed, interventions are commonly designed without consideration of the type of intervention likely to be effective, rationale for choice of interventions, the full range of possible influences on behaviour, and understanding the target behaviour [Davies et al., 2010; van Bokhervan et al., 2003]. Many frameworks exist but have not been intended for- or been tested in healthcare settings. The BCW framework (Figure 2) consists of the COM-B model at the core with its three critical components; surrounding this are nine intervention functions and seven policy categories that aim to address any COM-B components identified as deficient. Thus, each domain of TDF or COM-B can be used with the BCW to identify BCTs and design and refine interventions (Figure 2). The eight step process outlined by Michie et al. [2011] standardises a comprehensive approach to intervention development as follows: (1) define the problem in behavioural terms; (2) select the target behavior(s) most likely to bring about change to address the problem, (3) specify the target behavior in as much detail as possible, (4) identify what needs to shift in order to achieve the target behavior, (5) identify intervention functions, (6) identify policy categories, (7) identify behavioural change techniques, and (8) identify mode of delivery. In a further step APEASE criteria [Michie et al., 2014] can be used to refine or select BCTs and their delivery, for an intervention, by considering a wider context of affordability, practicability, cost-effectiveness, acceptability, safety and equity; in three included studies [Smits et al., 2018; Riordan et al., 2010; Moise 2020] this was undertaken via consensus methods and evidence review. Although now popular in healthcare research (and elsewhere) for its comprehensive and practical approach to intervention design, limitations have been acknowledged, in terms of over simplifying human behaviour, lack of stakeholder involvement including the target population, vagueness where judgement is required for decision making, and the requirement to understand psychological processes [O’Cathain et al., 2019].

**Figure 2.**
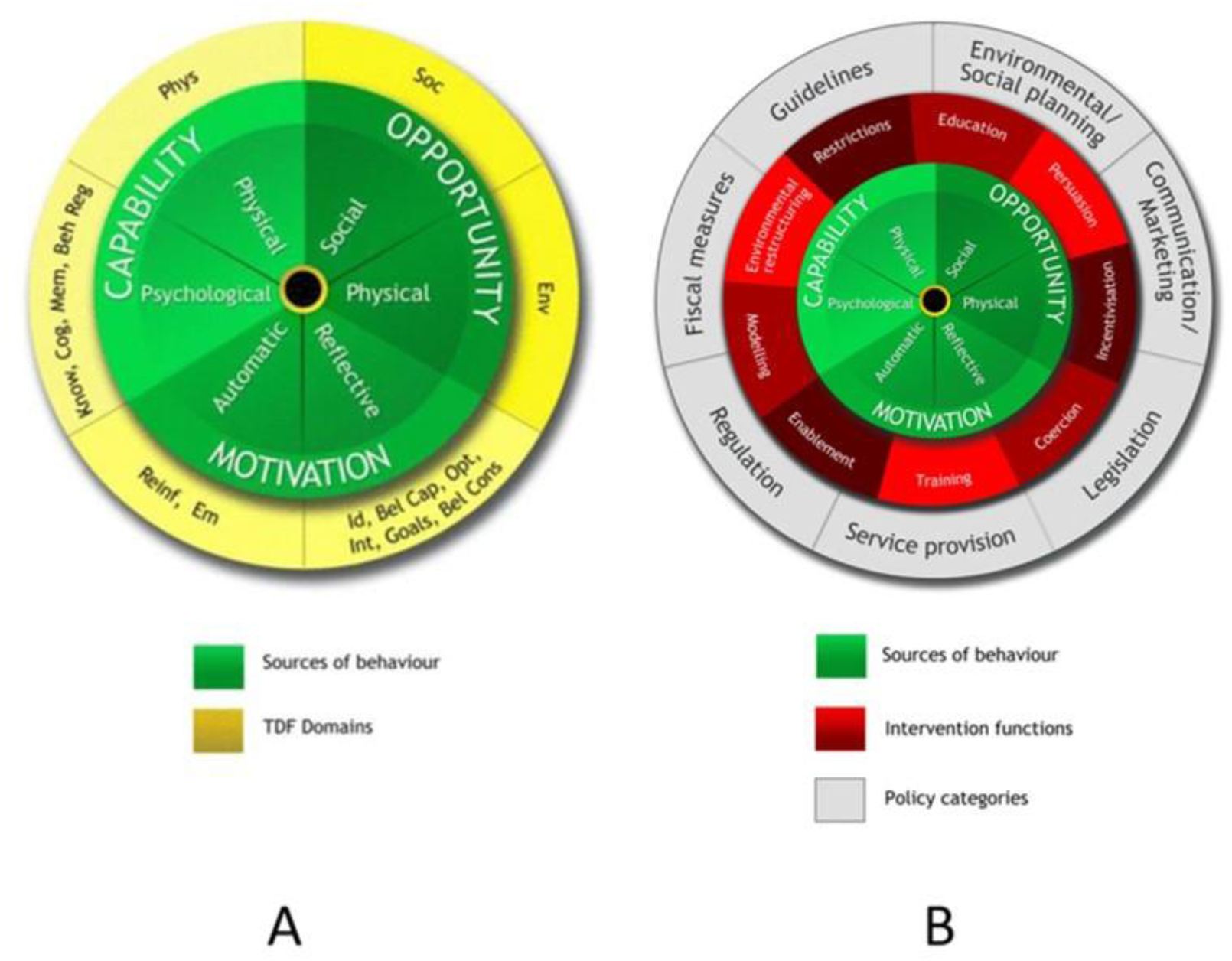
The behaviour change wheel (BCW): a new method for characterising and designing behaviour change interventions. A: Mapping between theoretical domains framework (TDF) and the COM-B model, frameworks for understanding behaviour and analysing relevant qualitative data; B=The BCW. Taken from Atkins et al. [2017] and Michie et al. [2011] respectively. Soc=social influences; Env=environmental context and resources; Id=social/professional role and identity; Bel Cap=beliefs about capabilities; Opt=optimism; Int=intentions; Bel Cons=beliefs about consequences; Reinf=reinforcement; Em=emotion; Know=knowledge; Cog=cognitive and interpersonal skills; Mem=memory, attention and decision processes; Beh Reg= behavioural regulation; Phys=physical skills.

Although no specific intervention or technique is recommended for use in patients with asthma and work-related asthma, there are some general and transferable insights into BCI design from a recent systematic review by Mather et al. [2022], focused on barriers and enablers of behaviour change by primary care HCPs. The authors reported that HCPs commonly perceive those in the ‘capability’ and ‘opportunity’ domains of COM-B, which are linked with interventions related to education, training, restriction, environmental re-structuring and enablement; these results are therefore in keeping with those of the only full-text study focused on the area of guidelines for unspecified chronic diseases, by Kredo et al. [2018], which identified high priority functions relating to education, training and enablement. A specific BCI for use with patients at risk of work-related asthma could be developed using a standardised and establish BC methodology, taking account of local variation in employment, industry, and causative exposures.

## Data Availability

All data produced in the present study are available upon reasonable request to the authors

