## Supplementary material for "Could behaviour change techniques be used to address under-recognition of work-related asthma in primary care? A systematic review"

<u>SUPPPMENTARY MATERIAL</u>

| Search terms and number of hits | | |
|---|---|---|
| 1 | (behaviour change or behavior change or BCI or behavioural change or behavioral change).ab. | 90376 |
| 2 | (primary care or general practice).ab. | 385243 |
| 3 | (diagnosis or identif$).ab. | 13222849 |
| 4 | 1 and 2 and 3 | 1598 |
| 5 | remove duplicates from 4 | 917 |
| 6 | limit 5 to abstracts | 916 |
| 7 | limit 6 to human | 817 |

**Supplementary Table 1. Literature search terms and number of hits.**

| **Rationale for exclusion** | **Explanatory notes** |
|---|---|
| Wrong study design | All protocols (study not yet been done or no results given), reviews (systematic or narrative), commentary or opinion pieces, theses, dissertations, book chapters, books, cross-sectional surveys.<br><br>Note, some protocols contain the detail on the development of the BCI, and these have been considered for inclusion on that basis. |
| Wrong populations | All studies undertaken in populations other than primary care (eg. acute medical admissions units, tertiary services, emergency department etc.). All studies that examine behaviour change relating to treatment or disease outcomes rather than diagnosis of disease. Research that evaluated or developed interventions for behaviour change related to adherence to screening programmes were considered for inclusion, but not development of screening programmes per se. |
| Wrong outcome measurements | Outcomes of studies that did not include either development of a BCI **or** evaluation of its effectiveness in practice (implementation), or both. |

**Supplementary Table 2. Rationales for study exclusion. BCI=behaviour change intervention.**

| Data variable | Explanatory notes |
|---|---|
| Authors | First author, et al. |
| Year | |
| Rationale | Broad aim(s) of study; development of BCI or evaluation, or both |
| Primary methodology | Eg. qualitative, RCT, quasi-RCT, quality improvement design, multi-component |
| Populations | Target population (eg. patients, HCPs, both); disease area of interest (eg. diabetes, epilepsy, asthma) |
| Structure and function of intervention (development and/or evaluation) | Narrative description of structure and focus of BCI (eg. multi-component training and feedback, single component audit/feedback tool) |
| Theoretical framework (development) | Any described theoretical basis described for BCI design and development |
| Comparator groups (evaluation) | Observational or experimental study designs, description of comparators, and whether any clustering, randomisation or blinding was undertaken |
| Outcome measures (evaluation) | Related to effectiveness or utility; any effect measures given (eg. proportions, risk ratios, prevalence ratios, incidence rate ratios) with confidence intervals and statistical measures of significance |
| Notes | Open comments to aid interpretation and writing |

**Supplementary Table 3. Data fields in data collection template. BCI=behaviour change intervention; RCT=randomised controlled trial; HCP=healthcare professional.**

**Supplementary material: Quality assessments**

Key

| | |
|---|---|
| Yes | (green) |
| No | (red) |
| Unclear | (amber) |
| Not applicable | |

| Studies where randomised controlled trials (RCT) performed | | | | |
|---|---|---|---|---|
| **Quality/bias assessor** | | **Papers** | | |
| | | **RCTs with cluster randomisation** | | **RCT with participant randomisation** |
| | | Kronish et al. (2022) [Conference abstract only] | Rubenstein et al. (2011) | Larkey et al. (2015) |
| Selection and allocation | Was true randomisation used to assign participants to treatment groups? | | | |
| | Was allocation to treatment groups concealed? | | | |
| | Were treatment groups similar at the baseline? | | | |
| Administration of intervention | Were participants blind to treatment assignment? | | | |
| | Were those delivering the treatment blind to treatment assignment? | | | |
| | Were treatment groups treated identically other than the intervention of interest? | | | |
| | Were outcome assessors blind to treatment assignment? | | | |
| | Were outcomes measured in the same way for treatment groups? | | | |
| | Were outcomes measured in a reliable way | | | |
| Participant retention | Was follow up complete and if not, were differences between groups in terms of their follow up adequately described and analysed? | | | |

| | |
|---|---|
| Statistical conclusion | Were participants analysed in the groups to which they were randomized? |
| | Was appropriate statistical analysis used? |
| Trial design | Was the trial design appropriate and any deviations from the standard RCT design (individual randomization, parallel groups) accounted for in the conduct and analysis of the trial? |

**Supplementary Table 4. Critical appraisal and risk of bias for papers describing randomised controlled trials.**

| **Study with uncontrolled trial design** | |
|---|---|
| **Quality/bias assessor** | **Porcheret et al. (2018)** |
| Is it clear in the study what is the ‘cause’ and what is the ‘effect’ (i.e. there is no confusion about which variable comes first)? | |
| Were the participants included in any comparisons similar? | |
| Were the participants included in any comparisons receiving similar treatment/care, other than the exposure or intervention of interest? | |
| Was there a control group? | |
| Were there multiple measurements of the outcome both pre and post the intervention/exposure? | |
| Was follow up complete and if not, were differences between groups in terms of their follow up adequately described and analyzed? | |
| Were the outcomes of participants included in any comparisons measured in the same way? | |
| Were outcomes measured in a reliable way? | |
| Was appropriate statistical analysis used? | |

**Supplementary Table 5. Critical appraisal and risk of bias for uncontrolled trial design.**

.

| **Studies using employing predominantly qualitative methodology** | | | | | | | | | | |
|---|---|---|---|---|---|---|---|---|---|---|
| **Paper** | **Bias/Quality indicator** | | | | | | | | | |
| | 1. Is there congruity between the stated philosophical perspective and the research methodology? | 2. Is there congruity between the research methodology and the research question or objectives? | 3. Is there congruity between the research methodology and the methods used to collect data? | 4. Is there congruity between the research methodology and the representation and analysis of data? | 5. Is there congruity between the research methodology and the interpretation of results? | 6. Is there a statement locating the researcher culturally or theoretically ? | 7. Is the influence of the researcher on the research, and vice- versa, addressed? | 8. Are participants, and their voices, adequately represented? | 9. Is the research ethical according to current criteria or, for recent studies, and is there evidence of ethical approval by an appropriate body? | 10. Do the conclusions drawn in the research report flow from the analysis, or interpretation, of the data? |
| Kredo et al. (2018) | | | | | | | | | | |
| Leather et al. (2022) | | | | | | | | | | |

| |
|---|
| Payne and Hysong (2014)<br><br>[Conference abstract only] |
| Tuot et al. (2022) |

**Supplementary Table 6. Critical appraisal and risk of bias for qualitative studies as primary methodology.**

| Paper | Narrative assessment of behaviour change methodology |
| --- | --- |
| Smits et al. (2018) | Multi-component BCI methodology, aiming to refine an existing intervention. Components included evidence synthesis by SR and thematic analysis of qualitative data derived from focus groups. Methods outlined for both components though not in detail. Barriers and facilitators identified and mapped using a BCW/TDF framework, described in detail. |
| Lester et al. (2005) | Development of an educational BCI employing the MRC complex interventions framework. Multi-phase process including a literature review and data from focus groups, methods outlined briefly not in detail. Feedback gained from users of the intervention (GPs) via cross-sectional surveys after initial session and booster session. Questions referred to self-reported improvements in attitude, awareness, knowledge, and satisfaction though no outcome measures relevant here |
| Jinks et al. (2015) | Conference abstract for co-design of an intervention employing a multi-component BCI. Components listed and included evidence synthesis (nature unclear), community of practice and qualitative focus group with practice nurses. No component methods described in detail here, though data mapped using a BCW/TDF framework. |
| Bravington et al. (2022) | Secondary coding of a qualitative data set (original study methods described elsewhere in detail) to identify barriers and facilitators for cervical screening, then employing a TDF framework in order to develop BCIs. Re-coding and categorisation methods described and referenced in this paper in detail. Qualitative data from service-user and HCP focus groups used to guide written content of intervention, described in detail. |
| Moise et al. (2020) | A protocol for a RCT for a BCI aimed to increase guideline uptake, though design and refinement of the BCI is described is detailed in the first part of the paper. A multi-component BCI methodology with a BCW/TDF framework described here in some detail. Qualitative focus group aspects exploring barriers, published in their entirety in a separate paper, and results summarised here in a table. Mapping of barriers to functions via the BCW undertaken by the research team and summarised in tables. Third aspects described were key informant interviews for feasibility, with methods in some detail. |
| Porcheret et al. (2014) | A protocol for a RCT for a BCI aimed to improve effectiveness of primary care consultations. A multi-component BCI methodology is described and draws on 4 theoretical frameworks (implementation of change model, TDF, theoretical mapping of behavioural determinants to BCTs (after Michie et al.), and principles of adult learning). There is outline on the methods employed eg. in conducting advisory groups, feasibility discussions, though results are summarised. |
| Riordan et al. (2020) | Development of a multi-component BCI aimed at HCPs and patients with type 2 diabetes, for improvement of diabetic retinal screening uptake. Undertaken through a multi-step process comprising initial audit of existing screening behaviour, re-analysis of existing qualitative data to identify barriers and enablers for HCPs and patients, identification of BCTs through mapping to TDF domains, refinement of final BCI by feasibility and usability testing via consensus group meetings with HCPs and patients and |

| | |
|---|---|
| | applying APEASE criteria. Each step described in methods section. Qualitative elements (consensus group work and exploration of barriers and enablers) described in detail but no detail on reflexivity. |
| Smith et al. (2012) | Iterative development of complex healthcare intervention (BCI) according to the MRC complex interventions framework. Each phase described in turn in detail, with results summarised. BCI refinement included qualitative focus groups methods outlined, and results given in some detail in main text. |
| Toh et al. (2016) | Conference abstract for a multi-component BCI aimed at pharmacists undertaking clinical assessments, developed using BCW theory and thematic analysis of interviews with patients and HCP stakeholders. Detail is lacking being a conference abstract, but sampling method is specified and technique stated (thematic analysis). However detail on reflexivity, ethical permissions, philosophical perspective is missing. Barriers identified from the thematic analysis is stated but precisely how the BCW was used to frame these is unclear. The barriers are described in the results briefly, as are the intervention functions, but any selection or refinement process is missing. |
| Tuot et al. (2020) | A protocol for an uncontrolled trial of a clinical toolkit (BCI) for case finding kidney disease (as well as a patient facing risk calculator), that also describes development and refinement of the BCI, using a BCW/TDF methodology. Early stage included qualitative interviews with patients and HCPs, though no detail on methods given, though interview schedule attached in supplement. Barriers and enablers are summarised in a table. |

**Supplementary Table 7. Summary of narrative assessment of quality for studies using behaviour change methodologies, for which no standardised assessment tool exists.**
